# GLP-1 Receptor Agonists as a Potential Fifth Pillar of GDMT in HFrEF (NYHA II–IV): A Multicenter Real-World Propensity-Matched Analysis

**DOI:** 10.64898/2026.04.13.26350824

**Authors:** Osman Yousafzai, Kainat Kanwal, Frank Annie, Sarah Rinehart

## Abstract

**Background:** Despite widespread adoption of contemporary guideline-directed medical therapy (GDMT), patients with heart failure with reduced ejection fraction (HFrEF) continue to experience substantial residual morbidity and mortality. Glucagon-like peptide-1 receptor agonists (GLP-1RAs) have demonstrated cardiometabolic benefits in diabetes and obesity, but their role in HFrEF remains uncertain.

**Objectives:** To evaluate whether the addition of GLP-1RAs to optimized GDMT is associated with improved clinical outcomes in patients with HFrEF (NYHA class II–IV).

**Methods:** We conducted a retrospective, multicenter cohort study using the TriNetX Research Network. Adults (≥18 years) with HFrEF (LVEF ≤40%) receiving GDMT between January 2020 and October 2024 were included. Patients treated with GLP-1RAs were compared with those on GDMT alone. After 1:1 propensity score matching, 1,518 patients were included in each cohort. Outcomes over 2 years included all-cause mortality, major adverse cardiovascular events (MACE), critical care utilization, and acute kidney failure. Time-to-event analyses were performed using Kaplan–Meier methods and Cox proportional hazards models.

**Results:** In the matched cohort (mean age ∼63 years, ∼33% female), GLP-1RA use was associated with significantly lower all-cause mortality compared with GDMT alone (12.8% vs 23.8%; hazard ratio [HR] 0.48; 95% CI 0.40–0.57; p<0.001), corresponding to an absolute risk reduction of 11.0%. MACE was also reduced (35.8% vs 47.4%; HR 0.64; 95% CI 0.58–0.72; p<0.001). Additionally, GLP-1RA therapy was associated with lower critical care utilization (18.4% vs 28.9%; HR 0.55; 95% CI 0.47–0.64; p<0.001) and reduced acute kidney failure (29.2% vs 37.3%; HR 0.67; 95% CI 0.59–0.76; p<0.001). Rates of pancreatitis and substance-related disorders were low and not significantly different between groups.

**Conclusions:** Among patients with HFrEF receiving contemporary GDMT, adjunctive GLP-1RA therapy was associated with significant reductions in mortality, cardiovascular events, and healthcare utilization. These findings support the potential role of GLP-1RAs as a novel, mechanism-complementary therapy in HFrEF. Prospective randomized trials are needed to confirm these observations and determine whether GLP-1RAs should be incorporated as a fifth pillar of GDMT.

## INTRODUCTION

Worsening heart failure (WHF) defined by recurrent or escalating congestion and low-output symptoms despite previously stable therapy marks a pivotal transition in the natural history of chronic HF. It frequently necessitates emergency care, intravenous diuretics, or hospitalization ^(1,2).^ Each WHF episode compounds neurohormonal activation, subclinical myocardial injury, and end-organ dysfunction, leading to stepwise deterioration in survival and quality of life, underscoring the need for preventive, disease-modifying strategies beyond acute decongestion ^(3).^

Heart failure with reduced ejection fraction (HFrEF) remains a major public health burden despite contemporary therapy. In the United States, approximately 6.7 million adults live with heart failure, and the prevalence continues to rise, with substantial mortality and cost implications ^(4)^. Current guidelines endorse a foundational “quadruple therapy” consisting of an angiotensin receptor–neprilysin inhibitor (or ACEI/ARB where ARNI is not used), a β-blocker, a mineralocorticoid receptor antagonist, and a sodium–glucose cotransporter-2 inhibitor for patients with symptomatic HFrEF. This regimen has revolutionized management and improved survival ^(5,6)^. Nevertheless, even among patients optimized on quadruple therapy, residual risk persists, manifesting as recurrent hospitalizations, progressive ventricular remodeling, and cardiovascular death. Real-world and registry data confirm that despite broad implementation of GDMT, substantial residual morbidity and mortality remain, driven by incomplete neurohormonal suppression, persistent inflammation, metabolic dysregulation, and suboptimal drug titration ^(7)^. These residual risks highlight the need for additional, mechanism-complementary therapies targeting metabolic and inflammatory pathways not addressed by current GDMT.

Glucagon-like peptide-1 receptor agonists (GLP-1RAs) are incretin-based therapies widely prescribed for type 2 diabetes mellitus (T2DM) and obesity ^(8)^. Large cardiovascular outcome trials in T2DM including LEADER (liraglutide), SUSTAIN-6 (semaglutide), and REWIND (dulaglutide) demonstrated significant reductions in major adverse cardiovascular events (MACE) and favorable effects on weight, blood pressure, and lipid profiles ^(9–11)^. More recently, semaglutide reduced cardiovascular events in patients with overweight or obesity without diabetes (SELECT) and improved symptoms, physical limitations, and weight in HFpEF with obesity (STEP-HFpEF), reinforcing the ability of GLP-1RAs to remodel the cardiometabolic milieu that fuels HF progression ^(12,13)^. However, evidence in established HFrEF remains inconclusive. Small, randomized trials of liraglutide in recently hospitalized or stable HFrEF (FIGHT and LIVE) did not improve clinical stability or left ventricular function and were associated with higher heart rate and more cardiac adverse events ^(14,15)^. These findings predated universal adoption of today’s quadruple GDMT. Thus, whether GLP-1RAs provide incremental heart failure–specific benefit when layered on contemporary GDMT remains an important and open question.

Accordingly, we conducted a large, multicenter, propensity-matched, real-world analysis to evaluate whether the addition of GLP-1RAs to optimized GDMT in patients with HFrEF (NYHA II–IV) confers incremental benefit in reducing mortality and cardiovascular events. We hypothesized that GLP-1RAs may represent a potential fifth pillar of evidence-based therapy in HFrEF.

## METHODS

We conducted a multicenter, retrospective cohort study using the TriNetX Research Network, a federated platform that aggregates de-identified electronic health records from 112 academic and community healthcare organizations across the United States. Adults aged ≥18 years with a documented diagnosis of heart failure (ICD-10: I50.x or I50.82) and a left ventricular ejection fraction (LVEF) ≤40% were eligible for inclusion, with the most recent LVEF measurement used for cohort assignment. Patients were required to be receiving guideline-directed medical therapy (GDMT), defined in TriNetX by the presence of at least one HF-related medication, between January 1, 2020, and October 30, 2024. The exposure cohort consisted of new users of glucagon-like peptide-1 receptor agonists (GLP-1RAs), while the comparator cohort included patients meeting identical HF criteria but without any GLP-1RA prescriptions. Patients were excluded if they had evidence of a mechanical circulatory support device (ICD-10-PCS 02HA0RS or 02HA0QZ), heart transplant status, type 1 diabetes, pregnancy, bariatric surgery or bariatric surgery status, or active chemotherapy. These exclusions were applied identically to both cohorts, with the additional exclusion of any GLP-1RA exposure in the comparator group.

The index date was defined as the first date on which all inclusion criteria were simultaneously met, and outcomes were assessed beginning 1 day after the index event and continuing for 730 days. Propensity score matching (1:1) using greedy nearest-neighbor methods without replacement was performed to balance demographics, comorbidities, laboratory values, and medication profiles specified within TriNetX, yielding 1,518 well-matched patients in each cohort. Outcomes were predefined by TriNetX and included all-cause mortality, major adverse cardiovascular events (MACE: myocardial infarction, ischemic stroke, or death), all-cause hospitalizations, critical care utilization (ICU services), acute kidney failure, pancreatitis, alcohol use disorders, substance use disorders, and cocaine use disorders. Time-to-event analyses were conducted using Kaplan–Meier methods with log-rank testing, and hazard ratios (HRs) with 95% confidence intervals (CIs) were estimated using Cox proportional hazards regression, with censoring at last known clinical contact. All analyses were performed within the TriNetX Analytics Platform on November 25, 2025.

## RESULTS

After 1:1 propensity score matching, 1,518 patients remained in each cohort, with excellent covariate balance across demographic, clinical, laboratory, and medication characteristics. The matched population had a mean age of 63.8 ± 12.3 years in the GLP-1RA cohort and 62.6 ± 13.1 years in the comparator cohort, with approximately one-third female in both groups. Comorbidities including diabetes mellitus, chronic kidney disease, chronic obstructive pulmonary disease, and prior nicotine dependence were similar between cohorts. Median follow-up duration was 615 days for the GLP-1RA group and 523 days for the comparator group.

During follow-up, treatment with a GLP-1 receptor agonist was associated with a significantly lower risk of all-cause mortality compared with GDMT alone (12.8% [195 events] vs 23.8% [361 events]; hazard ratio [HR] 0.48; 95% confidence interval [CI] 0.40–0.57; p < 0.001). Survival at 730 days was 84.95% in the GLP-1RA cohort compared with 71.85% in the comparator cohort, reflecting an absolute mortality risk reduction of 11.0%.

The incidence of major adverse cardiovascular events (MACE) was similarly reduced in GLP-1RA–treated patients (35.8% [544 events] vs 47.4% [719 events]; HR 0.64; 95% CI 0.58–0.72; p < 0.001). Survival probability at two years was 59.7% for the GLP-1RA cohort and 46.5% for the comparator cohort, corresponding to a 24% relative risk reduction.

Critical care utilization was also lower among GLP-1RA users. ICU-level care occurred in 18.4% (280 events) of the GLP-1RA cohort compared with 28.9% (439 events) of the comparator group (HR 0.55; 95% CI 0.47–0.64; p < 0.001). Acute kidney failure rates were similarly reduced (29.2% vs 37.3%; HR 0.67; 95% CI 0.59–0.76; p < 0.001). Rates of pancreatitis were low in both groups (1.2% vs 1.4%) and not significantly different. Alcohol-related disorders, general substance use disorders, and cocaine use disorders were infrequent and showed no significant differences.

Overall, in this large, real-world propensity-matched cohort of patients with HFrEF receiving contemporary GDMT, adjunctive GLP-1RA therapy was associated with substantial reductions in all-cause mortality, MACE, ICU utilization, and acute kidney failure. These benefits were consistent and sustained across the two-year follow-up period, supporting the potential role of GLP-1RA therapy as an effective adjunctive treatment in high-risk patients with HFrEF.

## DISCUSSION

In this large, real-world, propensity-matched cohort of patients with HFrEF already receiving contemporary quadruple GDMT, adjunctive GLP-1 receptor agonist (GLP-1RA) therapy was associated with significant reductions in all-cause mortality, MACE, acute myocardial infarction, and critical care utilization, along with a modest decrease in ventricular arrhythmias.

Our results are consistent with the broader cardiovascular benefits of GLP-1RAs demonstrated in major outcome trials among patients with diabetes or obesity. In individuals with type 2 diabetes at high cardiovascular risk, LEADER (liraglutide), SUSTAIN-6 (semaglutide), and REWIND (dulaglutide) each showed significant reductions in MACE and all-cause mortality, accompanied by favorable effects on body weight, blood pressure, and lipid profiles ^(9–11)^. These benefits extended beyond glycemic control, underscoring the pleiotropic vascular and metabolic effects of GLP-1RAs. More recently, SELECT extended this paradigm to individuals with overweight or obesity without diabetes, demonstrating a 20% reduction in MACE with semaglutide ^(12)^. Similarly, STEP-HFpEF showed that semaglutide improved symptoms, physical limitations, and exercise capacity in patients with obesity-related HFpEF, positioning the class as a promising cardiometabolic therapy across the spectrum of heart failure phenotypes ^(13)^.

Evidence in HFrEF, however, has been mixed. The FIGHT and LIVE trials did not show clinical benefit of liraglutide in advanced HFrEF and raised concerns about potential adverse cardiac effects, including increased heart rate and arrhythmias ^(14,15)^. These trials, while foundational, were limited by small sample sizes, short follow-up, and the absence of background ARNI or SGLT2 inhibitor therapy, now standard components of GDMT. Our findings, derived from patients managed in the contemporary quadruple-therapy era, suggest that GLP-1RAs may provide additional benefit when integrated into optimized care, and that the unfavorable safety signal observed in earlier studies may not extend to broader, real-world populations.

Mechanistically, GLP-1RAs may attenuate HF progression through several complementary pathways, including improved myocardial glucose utilization, enhanced endothelial function, and reduced oxidative stress and inflammation via cyclic AMP– and PI3K-Akt–dependent signaling. These agents also promote weight reduction, decrease visceral adiposity, and improve insulin sensitivity, thereby reducing metabolic stress on the failing myocardium ^(8, 16-18)^. In addition, modest blood-pressure lowering, natriuresis, and anti-atherogenic vascular effects may contribute to the lower incidence of ischemic events and reduced need for critical care observed in our analysis. The modest but statistically significant reduction in ventricular arrhythmias could reflect improved metabolic stability and autonomic balance, although GLP-1RAs can slightly increase heart rate and should be used with caution in advanced HF ^(18)^.

Emerging experimental and translational data suggest that GLP-1RAs and SGLT2 inhibitors exert complementary effects on myocardial energetics, GLP-1RAs promoting glucose oxidation and SGLT2 inhibitors enhancing ketone utilization, which may underlie additive cardiometabolic benefits when combined ^(19–21)^. While definitive HFrEF-specific randomized trials of GLP-1RAs are still needed, this mechanistic framework aligns with the observed risk reductions in our study and supports the evolving view of heart failure as a systemic cardiometabolic disorder rather than a purely hemodynamic syndrome.

This study should be interpreted in light of several limitations. Its retrospective design precludes causal inference; residual confounding and misclassification from EHR/ICD coding are possible despite 1:1 matching. Adherence, dose, and duration of GLP-1RA were unavailable, and physiologic markers (e.g., NT-proBNP, remodeling indices) were inconsistently captured. Although we required a minimum exposure period, time-related biases (e.g., immortal-time bias) can arise in observational treatment comparisons and should be considered when interpreting effect sizes. Generalizability may be limited to U.S. health systems. These caveats underscore the need for adequately powered, contemporary randomized trials to determine whether GLP-1RAs merit formal inclusion as a fifth pillar of GDMT in HFrEF.

## CONCLUSION

Among patients with HFrEF on optimized GDMT, the addition of a GLP-1 receptor agonist was associated with lower risks of mortality, MACE, acute myocardial infarction, and critical care utilization, along with a modest reduction in ventricular arrhythmias. Randomized controlled trials specifically designed for HFrEF are needed to validate these real-world findings and clarify whether GLP-1RAs merit inclusion as a fifth pillar of guideline-directed therapy.

## Data Availability

None

## Conflict of Interest Statement

Osman Yousafzai, MD, Kainat Kanwal, MD, Frank Annie, PhD, FACC -None

Sarah Rinehart, MD FACC – Speaks for the Company Heartflow

---

**Table 1:** Baseline Characteristics of both cohorts before and after Matching.

| Characteristic | Before Matching<br>GLP- | Before Matching<br>No-GLP- | p-value | SMD | After Matching<br>GLP- | After Matching<br>No-GLP- | p-value | SMD |
| --- | --- | --- | --- | --- | --- | --- | --- | --- |

|  | **1RA**<br>**(n=2,825)** | **1RA**<br>**(n=53,826)** |  |  | **1RA**<br>**(n=1,518)** | **1RA**<br>**(n=1,518)** |  |  |
| --- | --- | --- | --- | --- | --- | --- | --- | --- |
| **Age, mean ± SD** | 64.0 ± 12.1 | 69.3 ± 14.1 | <0.001 | 0.403 | 63.8 ± 12.3 | 62.6 ± 13.1 | 0.011 | 0.092 |
| **Female (%)** | 33.0% | 31.4% | 0.086 | 0.033 | 32.8% | 31.5% | 0.437 | 0.028 |
| **Male (%)** | 67.0% | 68.5% | 0.090 | 0.033 | 67.2% | 68.5% | 0.437 | 0.028 |
| **White (%)** | 68.8% | 71.3% | 0.003 | 0.056 | 68.5% | 66.6% | 0.261 | 0.041 |
| **Black (%)** | 23.5% | 21.6% | 0.014 | 0.047 | 24.6% | 25.8% | 0.452 | 0.027 |
| **Asian (%)** | 2.0% | 2.2% | 0.532 | 0.012 | 1.8% | 2.4% | 0.312 | 0.037 |
| **Hispanic/Latino (%)** | 5.8% | 5.6% | 0.637 | 0.009 | 6.4% | 6.9% | 0.560 | 0.021 |
| **Not Hispanic/Latino (%)** | 84.1% | 87.1% | <0.001 | 0.085 | 83.7% | 81.6% | 0.114 | 0.057 |
| **Unknown Ethnicity (%)** | 10.1% | 7.3% | <0.001 | 0.099 | 9.9% | 11.5% | 0.142 | 0.053 |
| **COPD (%)** | 25.2% | 25.0% | 0.814 | 0.005 | 25.9% | 26.2% | 0.869 | 0.006 |
| **Atherosclerotic Heart Disease (%)** | 74.5% | 61.9% | <0.001 | 0.272 | 74.2% | 75.0% | 0.588 | 0.020 |
| **Diabetes mellitus (%)** | 91.4% | 43.9% | <0.001 | 1.180 | 87.4% | 91.2% | 0.001 | 0.122 |
| **History of Nicotine Dependence (%)** | 48.7% | 38.7% | <0.001 | 0.203 | 47.6% | 47.8% | 0.913 | 0.004 |
| **Chronic Kidney Disease (%)** | 49.6% | 41.2% | <0.001 | 0.170 | 48.2% | 49.6% | 0.446 | 0.028 |
| **Alcohol Abuse (%)** | 5.8% | 8.1% | <0.001 | 0.089 | 6.9% | 6.1% | 0.378 | 0.032 |
| **HbA1c, mean ± SD** | 8.0 ± 2.0 | 6.5 ± 1.6 | <0.001 | 0.785 | 8.0 ± 2.1 | 7.4 ± 2.0 | <0.001 | 0.266 |
| **ACE inhibitors (%)** |  |  |  |  |  |  |  |  |
| - Lisinopril | 60.4% | 39.8% | <0.001 | 0.421 | 58.6% | 61.5% | 0.103 | 0.059 |
| - Enalapril | 3.5% | 3.0% | 0.157 | 0.026 | 3.6% | 4.2% | 0.348 | 0.034 |
| - Captopril | 2.5% | 2.1% | 0.146 | 0.027 | 2.8% | 2.9% | 0.913 | 0.004 |
| - Ramipril | 3.2% | 1.9% | <0.001 | 0.084 | 2.8% | 2.2% | 0.299 | 0.038 |
| **ARB/Sacubitril-Valsartan (%)** |  |  |  |  |  |  |  |  |
| - Valsartan | 43.3% | 19.2% | <0.001 | 0.539 | 45.5% | 50.6% | 0.005 | 0.102 |
| - Sacubitril/Valsartan | 34.1% | 13.0% | <0.001 | 0.514 | 37.7% | 41.6% | 0.026 | 0.081 |
| - Losartan | 36.9% | 23.3% | <0.001 | 0.299 | 36.6% | 38.3% | 0.330 | 0.035 |
| **Beta-blockers (%)** |  |  |  |  |  |  |  |  |
| - Metoprolol | 76.0% | 60.8% | <0.001 | 0.333 | 77.5% | 79.7% | 0.133 | 0.055 |
| - Carvedilol | 47.6% | 33.1% | <0.001 | 0.299 | 48.0% | 51.3% | 0.075 | 0.065 |
| - Bisoprolol | 1.8% | 1.2% | 0.010 | 0.045 | 1.6% | 1.4% | 0.652 | 0.016 |
| **Mineralocorticoid Antagonists (%)** |  |  |  |  |  |  |  |  |
| - Spironolactone | 49.1% | 24.3% | <0.001 | 0.533 | 51.8% | 56.4% | 0.011 | 0.093 |
| - Eplerenone | 2.9% | 1.2% | <0.001 | 0.118 | 2.8% | 3.4% | 0.295 | 0.038 |
| **SGLT2 inhibitors (%)** |  |  |  |  |  |  |  |  |
| - Dapagliflozin | 23.7% | 4.7% | <0.001 | 0.565 | 22.7% | 24.8% | 0.186 | 0.048 |
| - Empagliflozin | 36.1% | 6.0% | <0.001 | 0.795 | 31.0% | 37.2% | <0.001 | 0.130 |
| **GLP-1 RAs (%)** |  |  |  |  |  |  |  |  |
| - Semaglutide | 25.9% | 0.1% | <0.001 | 0.833 | 5.9%* | 2.0%* | <0.001 | 0.200 |
| - Liraglutide | 17.5% | 0.5% | <0.001 | 0.625 | 13.6% | 13.9% | 0.792 | 0.010 |
| - Dulaglutide | 29.7% | 0.2% | <0.001 | 0.907 | 10.4% | 8.4% | 0.062 | 0.068 |
| - Tirzepatide | 3.3% | 0.1% | <0.001 | 0.250 | 1.6% | 1.6% | 1.000 | <0.001 |

**Table 2:** Outcomes in both groups after Propensity Matching.

| **Outcome** | **GLP-1RA +**<br>**GDMTn/N (%)** | **GDMT**<br>**Onlyn/N**<br>**(%)** | **Risk**<br>**Difference** | **Risk**<br>**Ratio**<br>**(RR)** | **Odds**<br>**Ratio**<br>**(OR)** | **Hazard**<br>**Ratio**<br>**(HR)** |
| --- | --- | --- | --- | --- | --- | --- |
| **All-cause**<br>**mortality** | 195 / 1,518<br>(12.8%) | 361 / 1,518<br>(23.8%) | −11.0% | 0.54 | 0.48 | **0.48**<br>(0.40–<br>0.57) |
| **Major adverse**<br>**cardiovascular**<br>**events**<br>**(MACE)** | 544 / 1,518<br>(35.8%) | 719 / 1,518<br>(47.4%) | −11.6% | 0.75 | 0.64 | **0.64**<br>(0.58–<br>0.72) |
| **Acute**<br>**myocardial**<br>**infarction** | 191 / 1,518<br>(12.6%) | 226 / 1,518<br>(14.9%) | −2.3% | 0.84 | 0.82 | — |
| **Critical care**<br>**utilization**<br>**(ICU/advanced**<br>**support)** | 280 / 1,518<br>(18.4%) | 439 / 1,518<br>(28.9%) | −10.5% | 0.64 | 0.55 | **0.55**<br>(0.47–<br>0.64) |
| **Acute kidney**<br>**failure** | 443 / 1,518<br>(29.2%) | 566 / 1,518<br>(37.3%) | −8.1% | 0.78 | 0.67 | **0.67**<br>(0.59–<br>0.76) |

## Figure Legend

**Figure 1.**
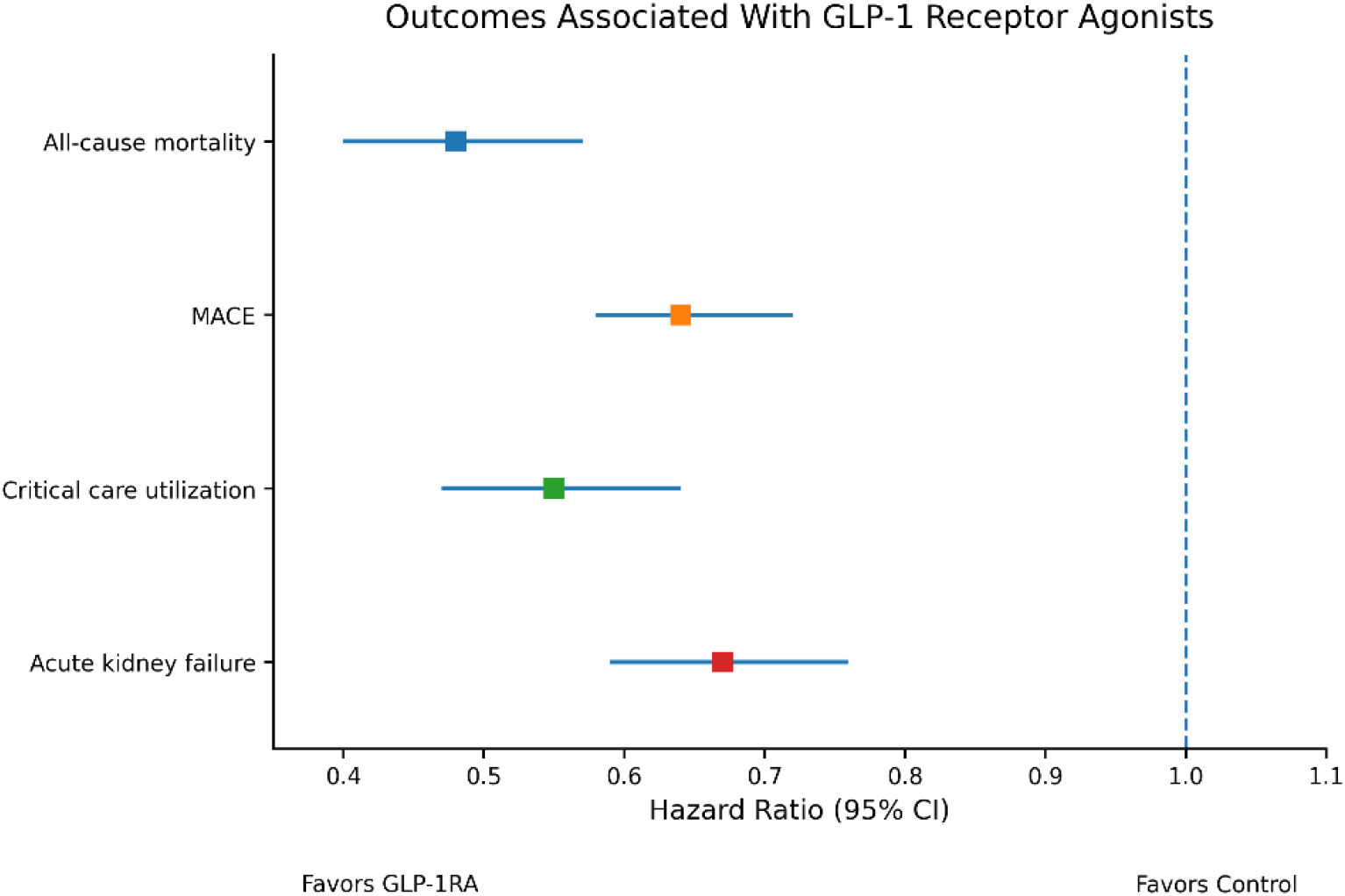
GLP-1 receptor agonists: outcome analysis

